# Correction of human forehead temperature variations measured by non-contact infrared thermometer

**DOI:** 10.1101/2020.12.04.20243923

**Authors:** Adrian Shajkofci

## Abstract

Elevated body temperature (fever) can be a common symptom of a medical condition, such as a viral or bacterial infection, including SARS-CoV-2 or influenza. Non-contact infrared thermometers are able to measure forehead temperature in a timely manner and were used to perform a fast fever screening in a population. However, forehead temperature measurements differ greatly from basal body temperatures, and are the target of massive perturbations from the environment. Here we gathered a dataset of N=18024 measurements using the same precision infrared sensor in different locations while tracking both outside temperature, room temperature, time of measurement, and identity. Herein, we propose a method able to extract and remove the influence of external perturbations and set the threshold for fever based on local statistics to 37.38 °C, after calibration and temperature correction. This method can help manufacturers and decision-makers to build and use more accurate tools so as to maximize both sensitivity and specificity of the screening protocol.

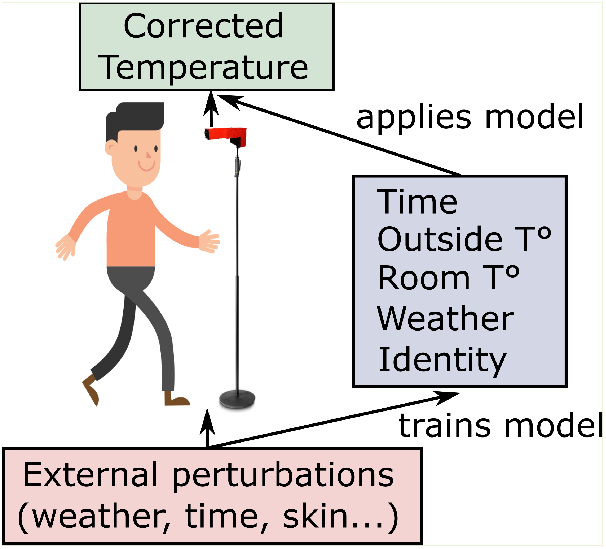

## I. Introduction

FEVER, also called pyrexia, is one of the usual clinical features that appears during the course of several infectious diseases, such as influenza or SARS-CoV-2 viral infections [1]. It reduces viral replication and is a straightforward marker of immune response [2]. Body temperature can be measured in numerous ways. The traditional method for such a measurement is using contact thermometers placed in the mouth, ear, armpit or rectum. Contact thermometers are measuring temperature using the conduction of heat to a thermocouple, a resistance temperature detector (RTD) or a thermistor through a metallic element. While RTDs are among the most precise temperature sensors available, the process of conduction is slow (around one minute) and the device needs extensive disinfection after use to prevent cross-contamination [3]. On the other hand, Non-contact infrared thermometers (NCITs) allow the temperature to be taken without contact and therefore do not require constant disinfection, take fast measurements (less than one second) and allow for a comfortable and much less intrusive measurement process. NCITs were extensively reviewed in [4] and validated for their use for fever symptoms detection.

NCITs can be classified into two categories. The first category comprises infrared cameras, also called thermal scanners. These devices operate like normal digital cameras, and capture a map of temperatures over a constant field-of-view (FOV). They are used to measure objects up to 10 meters in front of the camera. They focus heat radiation using coated optics made of germanium, zinc salts, or surface mirrors, towards a focal plane array (FPA) of heat-conductive pixels. The relative temperature of the FPA is then sequentially read by a bolometer that converts the temperature difference into resistance, further converted into electrical signals. Thermal imaging cameras require an extensive calibration using different sensitivity curves allocated to each pixel, their output being compared to the temperature of a black body. These devices usually need to be stabilized at a defined chip temperature [5]. Moreover, they are greatly dependent on the fluctuations of air temperature since the camera receives the radiation emitted from the targeted object, as well as heat coming from surrounding objects or the atmosphere [6]. According to Minkina and Klecha [7], the following parameters should be considered when calculating the temperature of the object: the emissivity of the object material (*ϵ*), the transmittance of the atmosphere, the distance between the object and the camera, the relative humidity, the atmospheric temperature *T*_*amb*_ and information about the surrounding heat sources (sun, halogen lamps, heated walls, etc.). These parameters are often difficult to gather and one must make approximations when designing a NCIT system. Moreover, these parameters can easily shift over time due to wind (or a draught), rain, or direct sunlight. Consequently, this type of sensor is known not to be accurate enough for medical temperature measurement. These devices are subject to an error of 0.5 ^*°*^C to 5 ^*°*^C when compared to contact thermometers, depending on the calibration modality [8]. The second type of NCIT is the family of thermopiles, mostly found in handheld forehead or tympanic thermometers. They receive the radiant energy released by the object over a constant FOV and convert it into an electrical signal (using the Seebeck effect), which is calibrated to the relative temperature of the object. Thermopiles are more accurate than infrared cameras since the ambient air flow is minimized due to the smaller distance between the sensor and the imaged object (1 cm to 20 cm) [9]. However, their accuracy drops drastically when used outside of their recommended range [10].

The accuracy of the measurement of human body temperature using NCITs is affected by individual factors, such as instrument placement, skin type and thickness, sweating, ethnicity, or body mass index [11], [12], as well as environmental factors such as outdoor temperature, wind, relative humidity, and level of direct sunlight. Indeed, these factors can either alter the temperature of the skin, or change the calibration parameters of the devices [13], [14].

During the 2020 COVID-19 pandemic outbreak, NCITs were used for fever symptom screening at quarantine stations. Their accuracy is discussed since skin temperature must become acclimatized after coming inside from the cold [15]. For example, in recent statistics from Kaohsiung Hospital [16], during the month of March, only 5 patients out of 40887 were identified with fever when the measurement was made outside. Yet, at a second indoor measurement checkpoint, further 37 people were identified with fever since their bodies have acclimatized to the indoor temperature.

Manufacturers and users of NCITs usually calibrate their sensors to the ambient temperature using blackbody calibration sources as a temperature of reference [17]. Nevertheless, this calibration method does not take into account the effect of the outside temperature on the human skin. Indeed, the skin acts as an insulative layer that, in the presence of cold, prevents heat loss in a both passive (fat layer) and active (vasoconstriction, arteriovenous shunts) manner [18]. For these reasons, without any correction, skin temperature becomes an erroneous marker for body temperature.

In our research, we consider the variation of human forehead temperature in different environments defined by separate locations, times of measurement within a day, outside temperatures, and room temperatures. We then examine if these perturbations influence the measurement accuracy in the context of infection symptom screenings. We believe that the analysis and corrections of external perturbations on human forehead temperature can be used to improve fever screening protocols by adapting the fever detection threshold to the variations. After having described the acquisition modalities in Section II-A, we design a statistical analysis of temperature variation of groups and individuals across time, ambient temperature, and meteorological conditions. Then, we propose in Section II-B a method to remove the effect of external perturbations. We publish the results in Section III. Finally, we discuss the results and suggest a threshold for fever detection in Section IV.

## II. Methods

### A. Acquisition modalities and sensor calibration

We collected the data from participating companies that used the infrared sensor module Coronasense (Coronasense, Martigny, Switzerland). The device embeds a thermopile-based sensor element (MLX90614-DCI, Melexis Technologies NV, Belgium) with a reported measurement accuracy of ≤ 0.2 ^*°*^C. The sensor chip is certified to comply with the ASSTM standard section 5.4 (Designation: E1965-98/2009)) - Standard Specification for Infrared Thermometers for Intermittent Determination of Patient Temperature. The devices were installed at the entrance of the buildings and measurements were taken approximately 30 seconds after entry. For each measurement, the subject was told to aim at a point 3 cm above the junction between the eyes, 10 cm away from the sensor. Exact distance between the device and the subject was controlled and recorded using a Light Detection And Ranging (LIDAR) detector. A burst of 10 measurements were taken at different positions on the forehead, and the maximal value was kept. For a subset of the data, we gathered an anonymous imprint of the subject’s identity, so we could track their temperature over time. For another subset of the data, we could also associate the measurements to the local meteorological conditions using OpenWeatherMap (OpenWeather Ltd., London, UK).

All sensors were factory calibrated using a black body and automatically calibrated themselves using the ambient air temperature. We adjusted the sensor output to the human body emissivity *ϵ* = 0.98 from Plank’s law [19]:

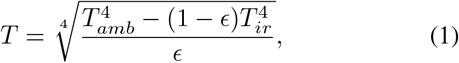

where *T*_*amb*_ and *T*_*ir*_ are the ambient temperature of the environment, and the mirrored temperature of the object, respectively. We added to every measurement an offset of 2.2 degrees in order to match with the average temperature obtained using a contact or a tympanic thermometer, as observed and concluded by [4], [20].

The acquired data is expressed as a mean and a variance. We calculated the coefficient of variance (CV) to determine the accuracy of the measurement. A CV under 5 % indicates a sufficient precision [21].

### B. Correction of environmental perturbations

We now turn to the task of measuring the correlation between the forehead temperature and environmental factors (inside ambient temperature, outside temperature and hours in the days) in the aim of removing the influence of these factors from the raw data. We model the relationship between the forehead temperature and the ambient and outside temperature data as a combination of affine functions *t*_*a*_(*t*) = *a*_*a*_*t* + *c*_*a*_ and *t*_*o*_(*t*) = *a*_*o*_*t* + *c*_*o*_ respectively. Furthermore, we model the relationship between the hours in a day and the forehead temperature as a second-degree polynomial function *t*_*p*_(*t*) = *t*^2^ + *t* + *c*_*p*_. We correct the temperature by subtracting to every point of data its corresponding point in the modeled curve or line and multiply, for every environmental perturbation, by the mean of the raw data.

In the same way, one can instead correct the threshold for fever detection by subtracting the offset between the regression curve and the acquired raw data to the fever threshold *t*_max_corr_. For example, to correct the threshold as a function of the outside temperature, we set:

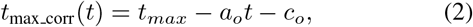

with *a*_*o*_ and *c*_*o*_ trained from the previous measurements using a least-squares fit.

When it is possible to track measurements of a unique individual over time, we propose a personalized threshold when the temperature measurements can be associated with an individual identifier, computed using the *K*-th last measurements of the specific person *i*:

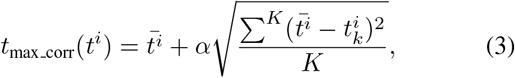

where 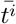 is the mean of *K* temperature acquisitions for the individual *i* and 0 *< α <* 5 a coefficient for the standard variation. We use *α* = 3 since it is the coefficient so that 99.7 % of the data remains detected as healthy under the normal approximation of the temperature distribution curve [22].

In the following section, for illustration purposes, instead of adapting the threshold dynamically for every data point, we adapted the individual temperature values against perturbations with a fixed threshold.

## III. Results

### A. Population analysis

We gathered *N* = 18024 measurements of temperature during a period of five weeks, using the same device, but in 6 different locations (see Figure 1 (a)). We computed statistics of the ensemble data for all locations as well as individual locations (Table I). We observed an average forehead temperature of 35.49 *±* 0.80 ^*°*^C. The mean and standard deviation of these measurements did not change significantly across different locations and among different populations (see Table I for locations A-C). We suppose that this consistency is the result of measuring among the population of the same country (French-speaking Switzerland), with similar meteorological conditions and ethnicity. The distribution of temperatures seems to be skewed toward low temperatures (see Figure 2 (a)).

**Fig. 1:**
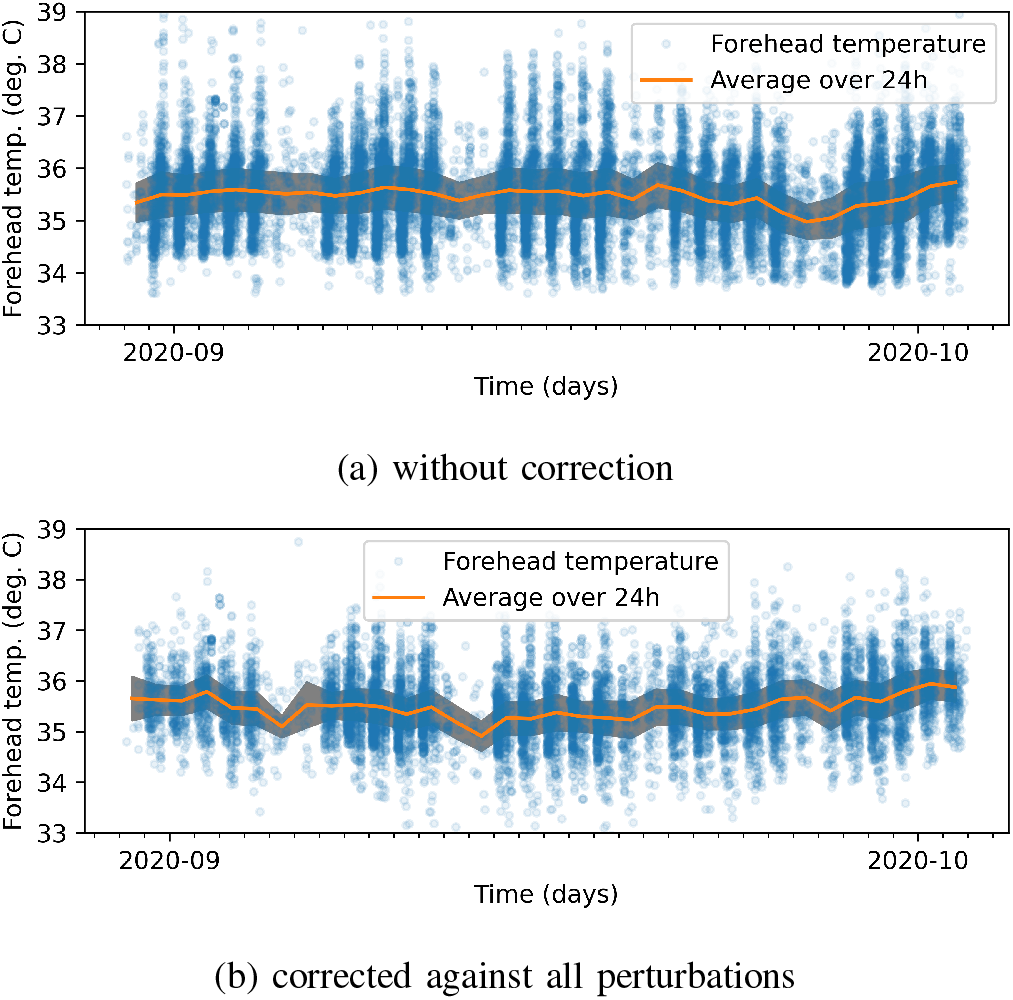
Dataset of forehead temperature measurements relatively to time, taken during a period of five weeks. Statistics are shown in Table I. (a) without correction (*N* = 18024) (b) with the corrections computed in Section II-B (ambient, outside and time corrections) (*N* = 10078)

**TABLE I:**
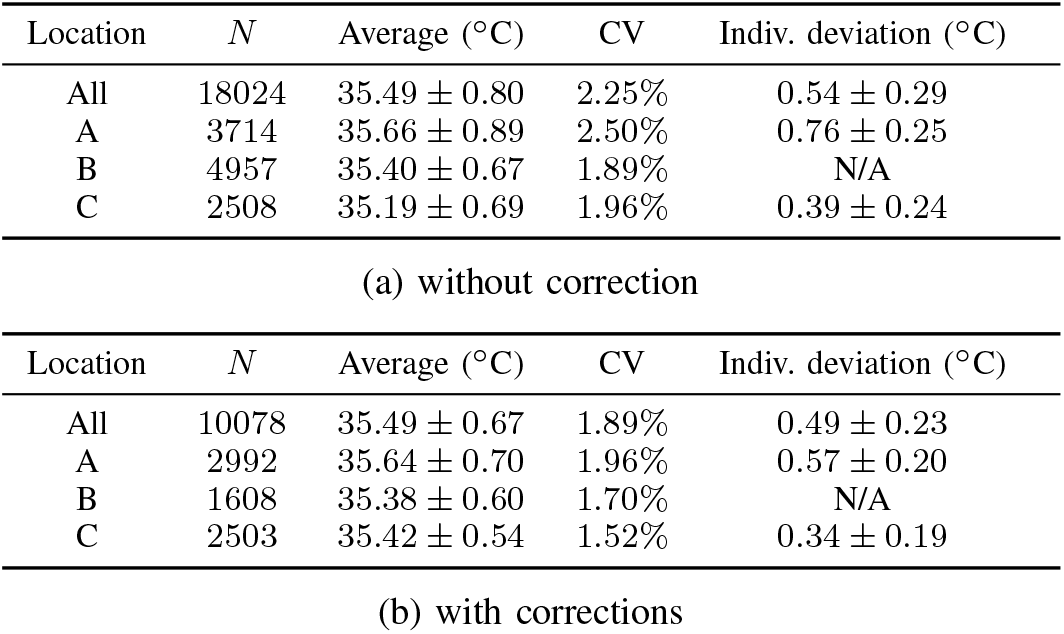
Statistics of the dataset of forehead temperature measurements (average, standard deviation, coefficient of variance (CV)) before and after correction. We also tracked temperatures of individuals over time and reported the mean of the deviation of temperature for one person over time. Because of the limited availability of meteorological data, we could compute such corrections on a subset of the original data.

**Fig. 2:**
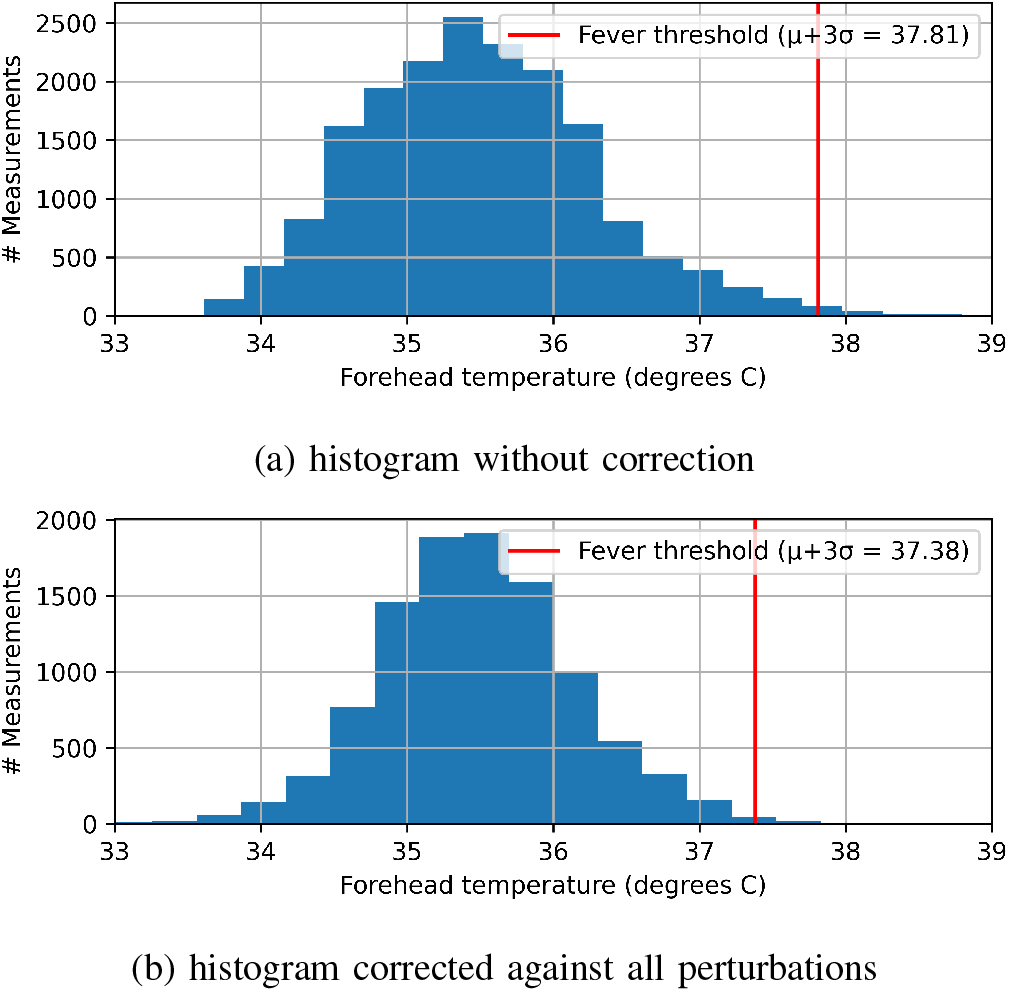
Histogram of forehead temperature measurements, taken during a period of five weeks. Statistics are shown in Table I. (a) without correction (*N* = 18024) (b) with the corrections computed in Section II-B (ambient, outside and time corrections (*N* = 10078))

### B. Tracking of individual temperature data

After having examined the data from an ensemble perspective, we now turn to the tracking of temperatures of individuals over time. For a subset of the data (*N* = 12882), we were able to gather individual anonymous tagging for every temperature measurement, along with all other features already analyzed in Section III-A. Results shown in Table I for variations of temperature of an individual over time indicates that the individual variation (0.54*±* 0.29 ^*°*^C) was lower than the average variation of temperature in a population (0.80 ^*°*^C).

### C. Correction of environmental perturbations

We were able to retrieve, on a subset of the dataset (*N* = 10078), the meteorological data of the exact measurement location at the time when the measurement was taken. We then trained the regression models from Section II-B. There was a significant correlation between outside temperature (*R*^2^ = 0.19) and forehead temperature in terms of Pearson correlation coefficient. Room temperature had a lower influence on the data (*R*^2^ = 0.12), since some measurements were taken when the individual entered a particular building. A similar correlation (*R*^2^ = 0.19) was found by comparing the measurements with their acquisition times within a 24-hour day. From the trained regression curves, we were able to subtract the differences representing the coupling of features for ambient, outside temperature and time of the day with the forehead temperature (*R*^2^ = 0) (see Figure 1 (b)). After correction, the variation of temperatures across both population and time were reduced, lowering the CV by 0.36 % (Table I (b)). The histogram of the corrected temperature density appeared to follow a Gaussian curve (Figure 2 (b)). The corrections were in the order of 0.02 *±* 0.51 ^*°*^C, with most of the corrections being in the [0 ^*°*^C; +0.5 ^*°*^C] interval (see Figure 3 (a)). By design, the corrections did not alter the mean of the temperature measurements. The corrections do not depend on the subject’s temperature but rather on the outside temperature (see Figure 3 (b)). For example, when the outside temperature was 5 ^*°*^C, the applied correction ranged from 0.65 ^*°*^C to 1.47 ^*°*^C.

**Fig. 3:**
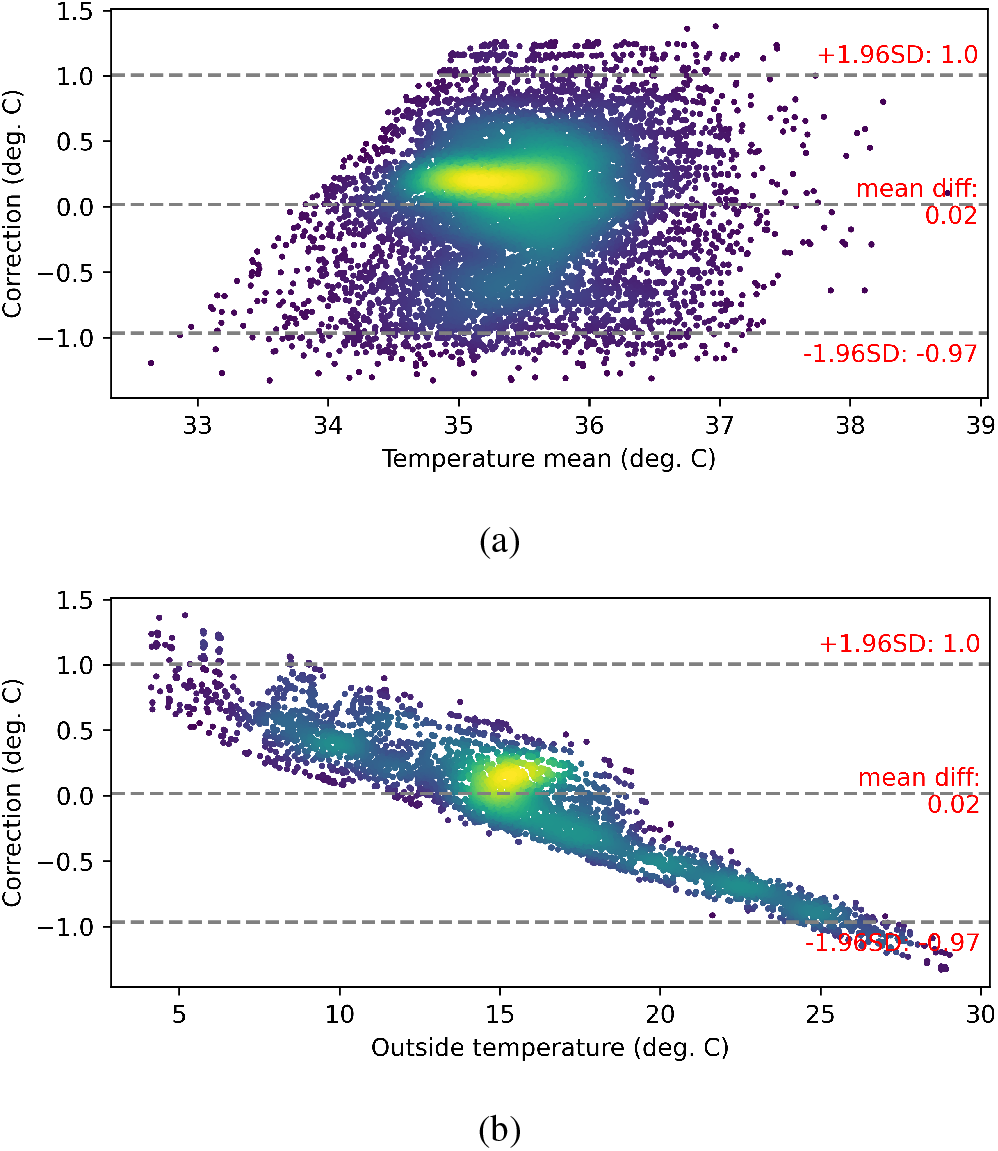
Bland-Altman plot of differences between uncorrected and corrected forehead temperature measurements (*N* = 10078). A negative value indicated that the corrected temperature is lower than the base temperature. The colors are the relative densities of measurements (the yellow color represent a higher density). (a) temperature correction values against mean temperature of corrected and uncorrected data points (b) temperature correction values against outside temperature

**Fig. 4:**
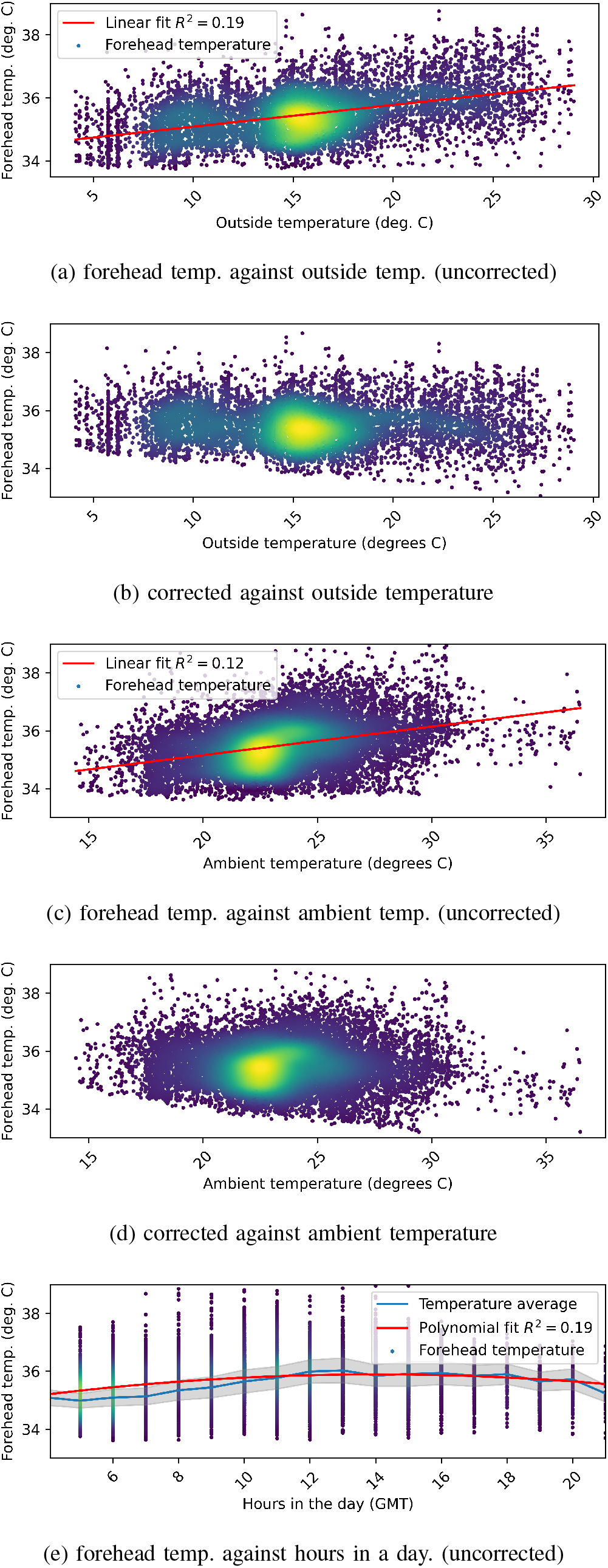
Dataset of forehead temperature measurements relatively to external perturbations ((a) outside temperature, (c) room temperature, (e) hour of measurement), taken during a period of five weeks, before ((a), (c), (e)) and after correction for environmental effects ((b), (d)). The number of measurements after outdoor temperature correction is smaller than without correction due to the availability of meteorological data. The color represents the relative point density (yellow is a higher density).

## IV. Discussion and conclusion

NCITs are remarkably noisy sources of data. Indeed, they exhibit variations due to different types of noise: intrinsic noise from the electronics; temperatures disparities due to sensor placement, skin type, skin color, and thickness, moisture, fat content; and finally, environmental conditions such as outside temperature, humidity, and sunlight exposure. Out of all these parameters, we chose to model the relationship between outside temperatures, ambient temperatures, acquisition time within the day and the measured forehead temperature as first and second-degrees polynomials. Using these models, we were able to subtract the influence of external perturbations and drastically reduce the co-linearity between these features. Thanks to the data analyzed in this study, we found out that the average forehead temperature was 35.49 *±* 0.80 ^*°*^C. This number was consistent across different places and matched to the values found in [23], [24], after proper scaling. Similarly to [4], we proposed a fever detection temperature threshold of the mean plus 3 times the standard deviation of the temperatures in our dataset (37.81 ^*°*^C with the offset we applied, or 35.68 ^*°*^C without calibration). After correction of external components, this threshold was reduced to 37.38 ^*°*^C, which is consistent with the results from [22]. Using the same calculation, this threshold could even be lowered due to the smaller deviation of measurements in specific environments (see Table I for Location B-C). Our results were not in agreement with the research by [4], who did not find a significant correlation between air temperature and body temperature. However, it appears that, in that study, the subjects were already acclimatized to a constant room temperature. We believe that, in a real-world case, temperature screening devices are installed at the entrance of a building and the security policy do not give the incoming people enough time to acclimatize inside the building before measuring their forehead temperature.

Furthermore, we found that the deviation of the forehead temperature of one subject over time is lower than the deviation of the mean temperature in a population, due to disparities between subjects. These findings confirm the idea that every individual is biologically distinct in terms of temperature and skin type due to demographic and physiological factors [25]. Consequently, the limit threshold for fever can be set individually as well. Personalized thresholds can be lower than the static threshold and might have a significant advantage over the latter for fever screening. Consequently, we propose a personalized fever detection threshold of the mean plus 3 times the standard deviation of the temperature record for a given subject, after correction for external perturbations. Such correlations between perturbations and the measured forehead temperature reveal that corrections to the temperature or fever detection threshold can lead to a more accurate recognition of high temperature symptoms. Eventually, even after corrections, our method does not take into account temperature perturbations caused by factors such as occasional sports, sweating and moisture on the skin. However, other data sources and modalities can be integrated to the computation, such as humidity measurements, tracking data or personal schedule of the subject to compensate for other perturbations. Our method might as well exaggerate false positives (type I error). For example, the alert might be triggered if the temperature outside is very low and the subject is warmed by an external source like a car heating. Hence, the challenge presented in [15], [16] is only partially settled. Finally, like with any fever screening method, we cannot exclude some form of examiner bias. Indeed, feverish subjects would be prone to either stay at home or try to avoid the screening process and therefore would not appear in the statistics.

In a time when disease outbreaks can spread very easily rapidly worldwide, fast disease screening solutions, such as fever screening, appear to be necessary. We hope that this research helps decision-makers and manufacturers to build more robust and intelligent tools that use multi-dimensional data in order to maximize both sensitivity and specificity of the screening protocol.

## Data Availability

Code and datasets are available online (GitHub).

https://github.com/ashajkofci/CorrectionForeheadTemperature

## V. Declarations

Code and datasets are available online: https://github.com/ashajkofci/ CorrectionForeheadTemperature. AS work for Coronasense, Martigny, Switzerland, who gathered and released the raw data.

## VI. Ethics statement

The authors used freely available anonymous data that was gathered with the consent of the owners of the measurement devices.

